# Impact of a novel mental health chatbot on depression knowledge among youth with HIV: A pilot study

**DOI:** 10.64898/2026.07.25.26358927

**Authors:** Jerome T. Galea, Carmen Contreras, Diego Vasquez, Neil Rupani, Karah Y. Greene, Milagros Tapia, Lenka Kolevic, Kristin Kosyluk, Molly F. Franke

## Abstract

**Introduction:** Adolescents living with HIV (ALWH) face disproportionately high rates of depression and anxiety, which can negatively affect antiretroviral adherence, viral suppression, and overall health outcomes. Despite the close relationship between mental wellness and HIV care, mental health services remain inadequately integrated into HIV programs, particularly in low- and middle-income countries. To address this gap, we developed EVA (*Educación, Vinculación y Autoayuda*), a mental health chatbot co-designed with ALWH to provide psychoeducation on depression and anxiety, teach self-help skills, and facilitate linkage to mental health resources. We evaluated EVA’s preliminary impact on depression knowledge and its feasibility and acceptability among ALWH in Lima, Peru.

**Methods:** During January–August 2024, ALWH in Lima, Peru completed baseline assessments, engaged independently with EVA for 20 minutes, and completed endline measures. The primary outcome was change in depression knowledge measured using the Adolescent Depression Knowledge Questionnaire (ADKQ). Secondary outcomes included acceptability, appropriateness, feasibility, intention to use and recommend the chatbot, and satisfaction with the chatbot’s features and content. Paired-samples t- tests were used to evaluate pre-post changes in depression knowledge.

**Results:** Fifty ALWH aged 11–19 years participated, among whom 70% reported greater-than-minimal depressive symptoms, 60% greater-than-minimal anxiety symptoms, and 92% moderate or high perceived stress. Following a single interaction with EVA, depression knowledge increased significantly, from a mean ADKQ score of 6.88 (SD=1.85) to 8.12 (SD=2.20), t(49) = -5.03, p<.001, representing a large effect size (d=1.74). The chatbot was rated highly for acceptability, appropriateness, and feasibility, and participants reported strong intentions to use and recommend it, as well as high satisfaction with its educational content, usability, and self-help resources.

**Conclusions:** A brief interaction with a mental health chatbot developed with and for ALWH in Peru significantly increased depression knowledge and was rated highly acceptable and feasible. Mental health chatbots may offer a low-cost, scalable approach to support mental health education and linkage to care within adolescent HIV services. Future longitudinal studies should assess sustained impact on mental health outcomes and help-seeking behavior; however, the benefits of chatbots should be weighed against their potential mental health risks.

## Introduction

Although the overall global HIV incidence and AIDS-related mortality declined significantly over the past two decades [1], for children and adolescents, these reductions have lagged behind those for adults. Approximately 2.4 million children and adolescents live with HIV (about 11% of the overall global HIV prevalence), accounting for 44% of new infections annually [2]. Distressingly, among the estimated 630,000 AIDS-related deaths in 2024, 14% were among children and youth aged 0-19 years [3].

Adolescence is a developmental period characterized by profound biological, psychological, and social transitions [4]. For adolescents living with HIV (ALWH), this period is often complicated by the burden of managing a chronic infection [5]. Mental distress among ALWH is common, especially depression, with a recent meta-analysis including N=2642 adolescents across 10 studies finding depression rates of 29.82% and 37.09% among youth aged 10-14 and 15-19 years, respectively [6]. Untreated depression can affect antiretroviral (ARV) adherence, compromise viral suppression, and worsen clinical outcomes [7-9]. Particularly for older adolescents, independently maintaining healthy behaviors like ARV adherence can be complicated by increased academic, social, romantic, and future-oriented demands [10, 11], including issues related to HIV disclosure and stigma [12, 13]. Despite clear evidence linking mental wellness to ARV adherence [14, 15], however, mental health services are frequently segregated from HIV prevention and care programs [16-19].

Many factors impact the integration of mental health and HIV care, especially in low- and middle-income countries (LMICs). Foremost among these are structural barriers, such as limited funding for mental health services and a critical shortage of trained psychiatric personnel [20]. These factors can limit people with HIV from accessing routine mental health services as part of their HIV care [21]. In practice, this often means only ALWH with severe psychological distress may be identified and linked to mental health services. This is a missed opportunity for early intervention to prevent or slow escalation of mental health symptoms and their impact on HIV. Previous work in Peru (an LMIC), where the HIV prevalence among adolescents aged 10-19 years has increased annually for the past two decades [22], is illustrative: among a cohort of 25 ALWH (mean age=18.9 years, range 15–21) with greater-than-minimal depressive symptoms, 92% (23/25) were not severe, and overall 12% (3/25) were referred to specialized mental health care [23]. Providing mental health support to these adolescents, especially education on anxiety and depression, self-help skills, and linkage to care, could be a scalable and economic way to promote mental health service integration with HIV care.

Chatbots (conversational tools that interact with users through text or speech in a manner that mimics human conversation to provide information) could play a role in addressing the mental health gap among ALWH. Chatbots can be accessed across various platforms such as SMS text messaging, WhatsApp, and Facebook Messenger without requiring additional software or apps, making them particularly useful in resource-limited environments. Use of chatbots in both consumer and healthcare has become widespread over the past decade due to their ability to provide information in a low-cost, scalable manner, particularly in settings with limited human resources [24, 25]. Regarding mental health specifically, chatbots have become increasingly prevalent and complex in their ability to deliver both information and therapeutic interventions [26, 27] though their use in LMIC, particularly for specific populations with unique needs, remains limited [28]. Nonetheless, in Peru during the COVID-19 pandemic, our team observed that ALWH possessed mobile devices and appreciated engaging with technology for information and communication [29], which led to the supposition that a chatbot customized for the unique needs of ALWH could help address the mental health care gap for this population.

Drawing on previous experience developing a sexual health chatbot for youth in the U.S. [30] and a health services access chatbot for Peruvian adults [31], we developed and pilot-tested a prototype mental health chatbot customized for ALWH aged 10-19 years in Peru. We hypothesized that a mental health chatbot customized to the unique needs of ALWH would increase depression knowledge and be feasible and acceptable.

## Methods

### Overview

We developed and pilot-tested a novel mental health chatbot for youth living with HIV in Lima, Peru. The chatbot had three core functions: providing psychoeducation on depression and anxiety; teaching mental health self-help skills; and providing linkage to existing mental health services. For the full study protocol, see [32]. The main outcome tested was a change in depression knowledge after a single exposure to the chatbot, and its acceptability and feasibility among ALWH were also assessed. The chatbot was developed in close collaboration with a Youth Advisory Board (YAB) comprised of N=6 youth aged 10-19 years living with HIV. The YAB also named the chatbot “EVA”, derived from the chatbot’s functions: ***E****ducación* (educate)*, **V**inculación* (linkage to care)*, **A**utoayuda* (self-help).

### Study Setting, Participants, and Procedures

A full description of EVA’s human-centered, multicomponent design phase leading to the present study is documented elsewhere [33]. All study procedures were conducted in Lima, Peru, over a period of approximately 18 months by the mental health program at the non-profit organization *Socios En Salud* (SES). Briefly, prior to the current study, EVA was developed in two phases: Phase I (months 0-6), qualitative data were collected from youth living with HIV, their caregivers, and HIV care professionals to understand their views on depression and their perceptions and preferences for a mental health chatbot tailored for youth with HIV [34, 35]. Phase II (months 7-12): Informed by Phase I data, EVA was designed and programmed using SmarBot360 [36], a user-friendly chatbot programming platform. Phase II involved an iterative, human-centered design process in close collaboration with the YAB, during which EVA’s content and features were determined, tested, critiqued, and refined [33].

Here, we focus on Phase III (months 13-18), during which EVA was pilot-tested in a controlled research setting among youth aged 10-19 years aware of their HIV diagnosis and naïve to the Phase I and II development processes described above (see: [32, 33, 35, 37]). Youth with acute emotional, physical, or social conditions were excluded from study participation and provided appropriate services. Phase III participants were recruited in coordination with the *Instituto Nacional de Salud del Niño*, the primary provider of pediatric HIV care in Peru; interested youth were referred to SES.

Study procedures were conducted in a private space at SES’ office; youth <18 years of age were accompanied by a guardian. Participants aged 18-19 years provided written informed consent, while youth <18 years old provided informed assent, with written informed consent from their guardian. Participants then completed a baseline assessment, after which they had 20 minutes of unstructured time to explore EVA individually. Finally, an endline assessment was administered. Participants received S/100.00 (about USD 25) for participation. A Peruvian Institutional Review Board approved the study (protocol N° 9651), which was ceded to by the University of South Florida (USF IRB study # 005124).

### Measures

Baseline and endline measures are detailed in the study protocol [32] and Table 1. In addition to a sociodemographic survey, we collected information on current depression and anxiety symptoms and perceived stress (Patient Health Questionnaire-9 [38, 39], Generalized Anxiety Disorder-7 [40]; Perceived Stress Scale [41]); mental health stigma and attitudes toward help seeking (The Self-Stigma of Seeking Help Scale [42], Attitudes Towards Mental Health Treatment [43]; and the acceptability and feasibility of the chatbot (Acceptability of Intervention, Intervention Appropriateness, and Feasibility of Intervention Measures [44]). Some instruments were modified (as noted in Table 1) at the YAB’s request to improve their suitability for youth respondents. The primary outcome of interest was change in depression knowledge after using the chatbot, measured using the Adolescent Depression Knowledge Questionnaire [45].

**Table 1:**
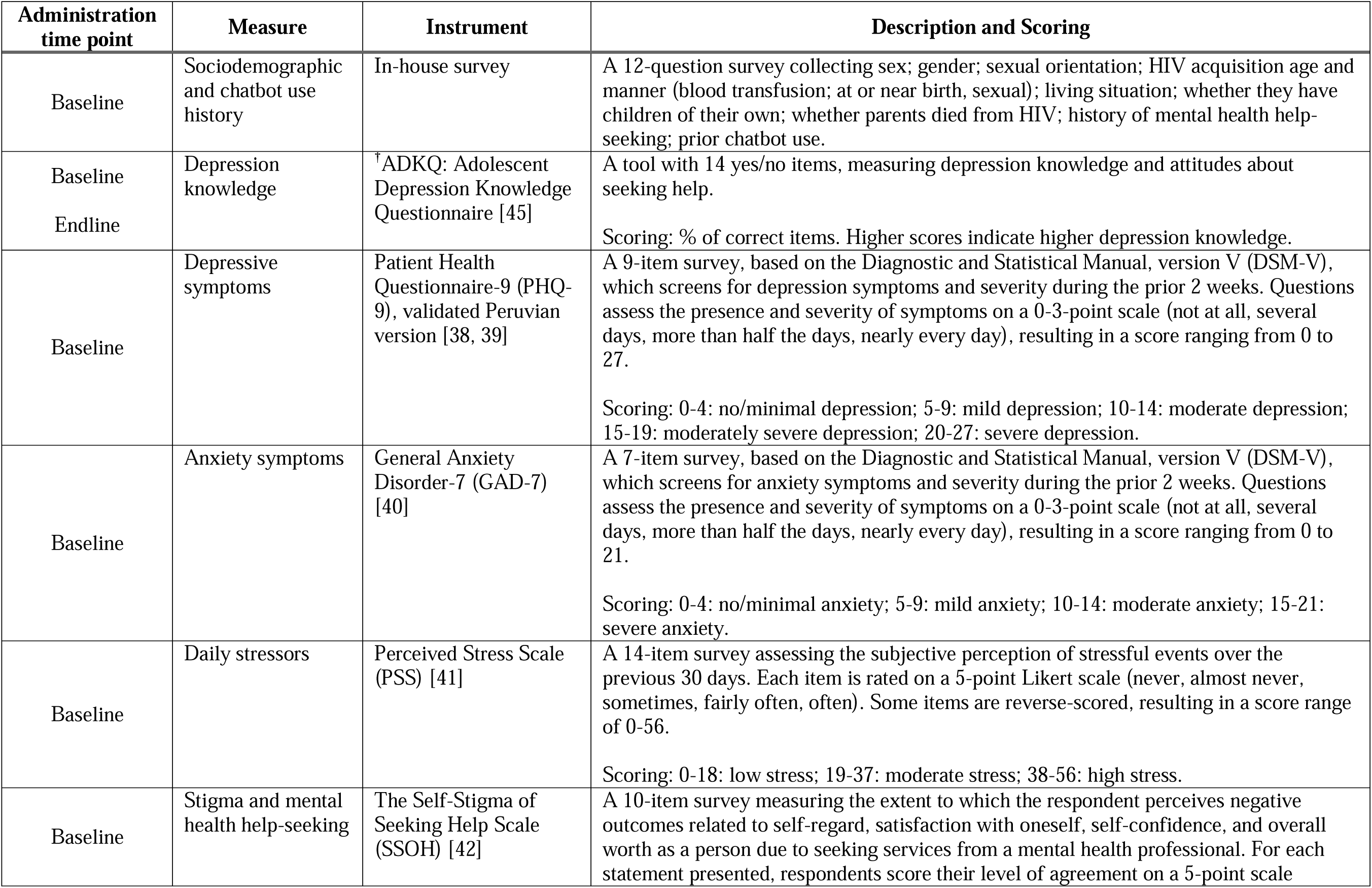
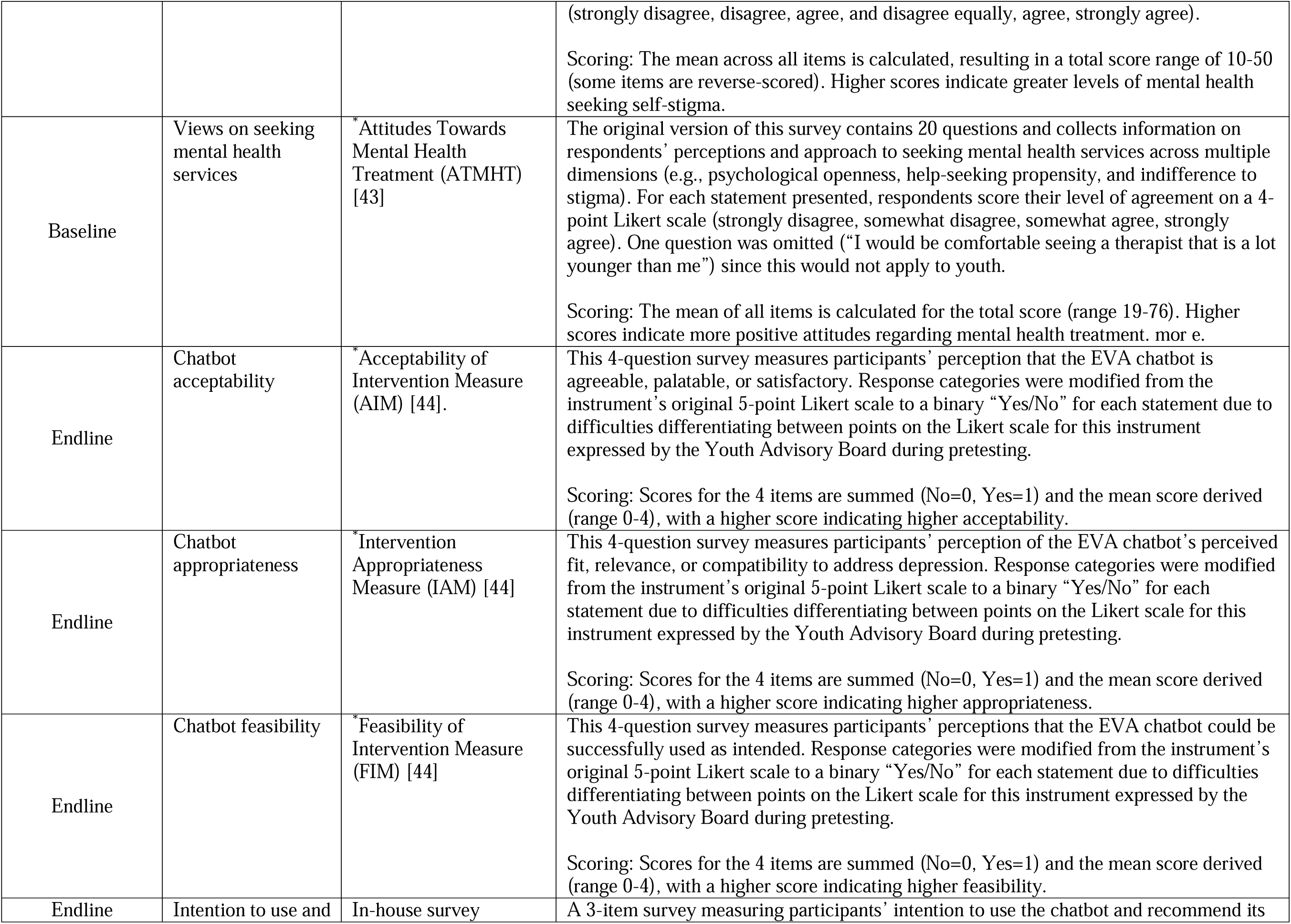
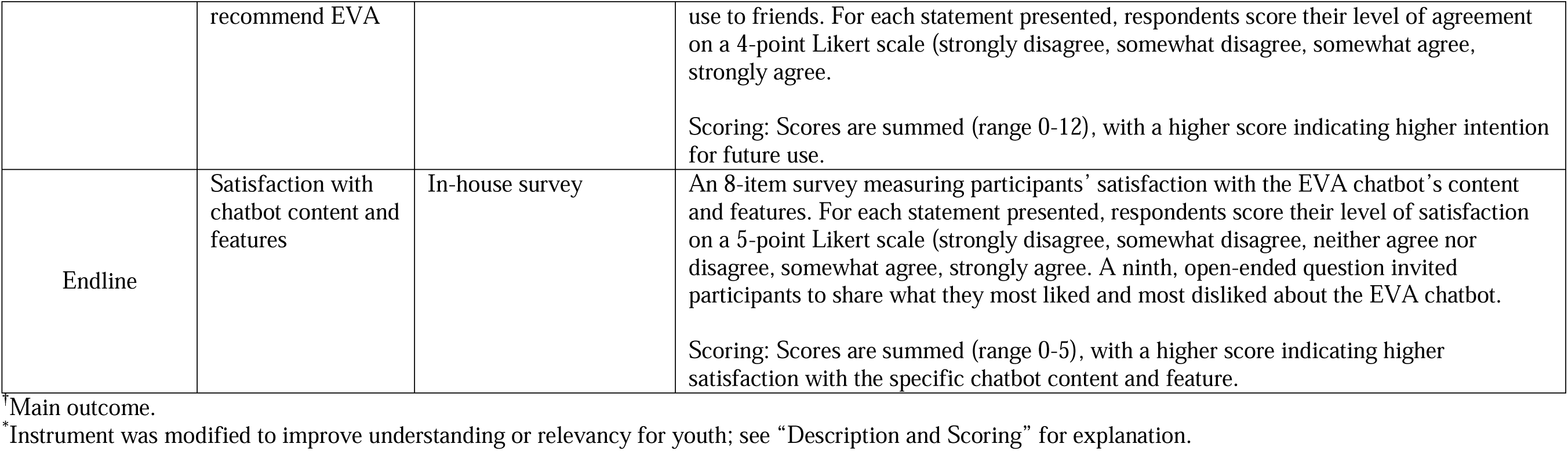
Measures and Instruments completed at baseline and endline, and item description.

### Sample Size, Data Processing and Statistical Analysis

An *a priori* power analysis was conducted using G*Power (version 3.1; Heinrich Heine University Düsseldorf) to determine the sample size needed to detect changes in depression knowledge, as measured by the ADKQ, following use of the EVA chatbot. The analysis was specified for a one-tailed paired-samples t-test (difference between two dependent means), assuming a medium effect size (d_z_=0.50), an alpha level of 0.05, and 95% statistical power. Results indicated that a minimum sample of 45 participants would be required to detect a significant increase in depression knowledge, if present. Data were collected on paper forms and entered into a secure, digital database. Descriptive statistics were generated based on each instrument’s scoring rules (Table 1).

## Results

### Sociodemographic characteristics

Fifty ALWH participated in the study, including n=11 (4 males, 7 females) aged 11-14 years and n=39 (27 males, 12 females) aged 15-19 years (overall mean =16.50 years [IQR=3]) (Table 2). Sixty-four percent (n=32) of all participants identified as heterosexual; 26% as homosexual (n=13); and 8% (n=4) as bisexual. Among the 11–14-year-olds, most (82%, n=9) acquired HIV at birth, while among 15–19-year-olds, most (69%, n=27) acquired HIV during sexual intercourse. Among all participants, 94% (n=47) reported current HIV antiretroviral use. Loss of at least one parent to HIV was more prevalent among 11–14-year-olds (64%, n=7) versus 15–19-year-olds (21%, n=8). Overall, most (59%, n=29) participants lived with at least one parent.

**Table 2:** Characteristics of ALWH participating in the chatbot study (N=50)

| Characteristic | 11-14 years old (n=11) |  | 15-19 years old (n=39) |  | Total (N=50) |  |
| --- | --- | --- | --- | --- | --- | --- |
|  | n | % | n | % | N | % |
| **Sex** |  |  |  |  |  |  |
| male | 4 | 36 | 27 | 69 | 31 | 62 |
| female | 7 | 64 | 12 | 31 | 19 | 38 |
| **Education** |  |  |  |  |  |  |
| only attending school | 10 | 91 | 19 | 49 | 29 | 58 |
| only working | 0 | 0 | 10 | 25 | 10 | 20 |
| working and attending school | 0 | 0 | 5 | 13 | 5 | 10 |
| other/no response | 1 | 9 | 5 | 13 | 6 | 12 |
| **Gender identity** |  |  |  |  |  |  |
| masculine | 4 | 36 | 23 | 59 | 27 | 54 |
| feminine | 7 | 64 | 12 | 31 | 19 | 38 |
| transgender | 0 | 0 | 2 | 5 | 2 | 4 |
|  | n | % | n | % | N | % |
| other/no response | 0 | 0 | 2 | 5 | 2 | 4 |
| **Sexual orientation** |  |  |  |  |  |  |
| heterosexual | 9 | 82 | 23 | 59 | 32 | 64 |
| homosexual | 0 | 0 | 13 | 33 | 13 | 26 |
| bisexual | 2 | 18 | 2 | 5 | 4 | 8 |
| other/no response | 0 | 0 | 1 | 3 | 1 | 2 |
| **HIV acquisition** |  |  |  |  |  |  |
| at birth | 9 | 82 | 12 | 31 | 21 | 42 |
| sexual | 2 | 18 | 27 | 69 | 29 | 58 |
| **Current ART use** |  |  |  |  |  |  |
| yes | 11 | 100 | 36 | 92 | 47 | 94 |
| no | 0 | 0 | 3 | 8 | 3 | 6 |
| **Pregnant or has children** | 0 | 0 | 5 | 13 | 5 | 10 |
| **Parents died from HIV** |  |  |  |  |  |  |
| at least one | 7 | 64 | 8 | 21 | 15 | 30 |
| neither | 4 | 36 | 31 | 79 | 35 | 70 |
| **Living situation** |  |  |  |  |  |  |
| alone | 0 | 0 | 3 | 8 | 3 | 6 |
| with one or both parents | 5 | 45 | 24 | 62 | 29 | 58 |
| with a non-parent family member | 5 | 45 | 4 | 10 | 9 | 18 |
| at a group home for children with HIV | 0 | 0 | 2 | 5 | 2 | 4 |
| with a friend | 0 | 0 | 5 | 13 | 5 | 10 |
| other/no response | 1 | 9 | 1 | 3 | 2 | 4 |

### Baseline depression and anxiety symptom prevalence and severity

Figure 1 displays the frequency and distribution of depression and anxiety symptom severity among participants. Greater than no/minimal depressive symptoms (PHQ-9 ≥ 5) were reported by 70% (35/50) of participants, of which 86% (30/35) were aged 15-19 years. Among participants with greater than minimal depressive symptoms, the distribution of PHQ-9 scores was: 40% (14/35) mild (PHQ-9 = 5-9); 29% (10/35) moderate (PHQ-9 = 10-14); 23% (8/35) moderately severe (PHQ-9 = 15-19); and severe 12% (3/35) (PHQ-9 = 20-27). All severe depression scores were among the 15-19-year-olds. Greater than no/minimal anxiety symptoms (GAD-7 ≥ 5) were reported by 60% (30/50) of participants, of which 83% (25/30) were aged 15-19 years. Among participants with greater-than-minimal anxiety symptoms, the distribution of GAD-7 scores was: 50% (15/30) mild (GAD-7 = 5-9); 37% (11/30) moderate (GAD-7 = 10-14); 13% (4/30) severe (GAD-7 = 15-21).

**Figure 1:**
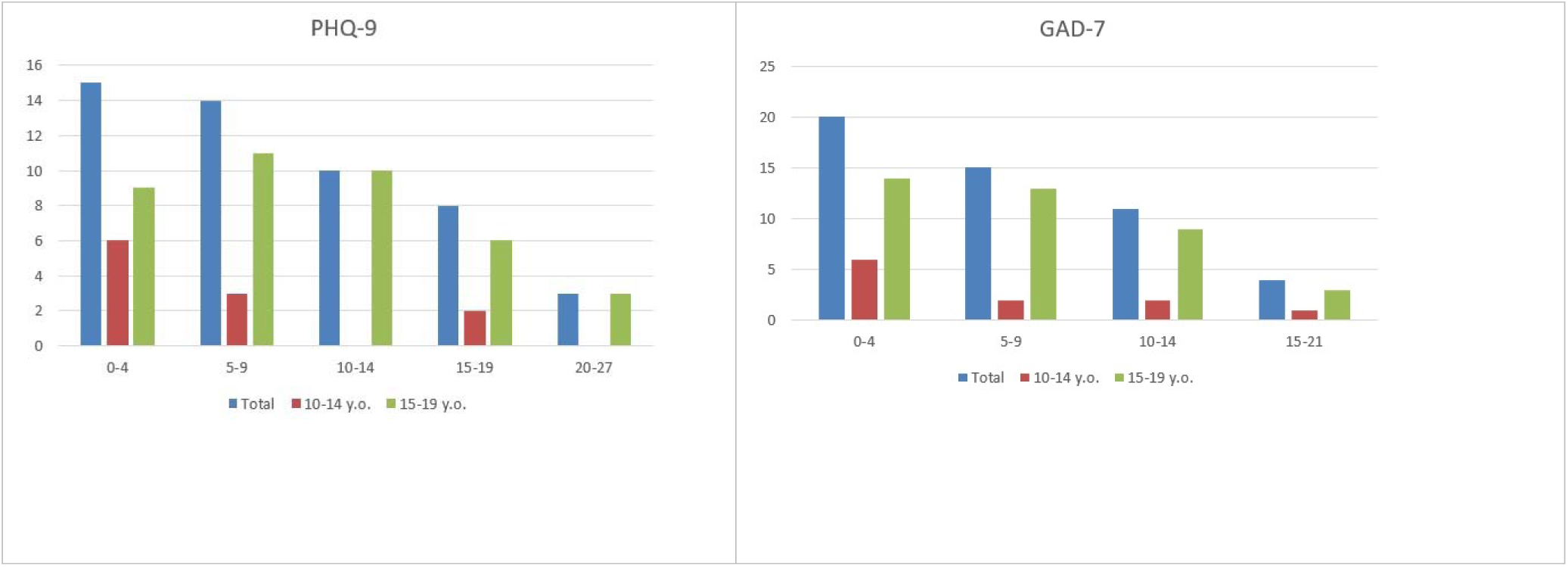
Baseline depression (PHQ-9) and anxiety (GAD-7) symptom prevalence and severity among N=50 ALWH participating in the EVA chatbot pilot study.

### Perceived stress and self-stigma of seeking help

Figure 2 displays the frequency and distribution of perceived stress severity among study participants. Among all participants, 8% (4/50) reported low stress (PSS = 0-18); 84% (42/50) reported moderate stress (PSS = 19-37); and 8% (4/50) reported high stress (PSS = 38-56). Participants aged 15-19 (N = 39) reported the highest frequency of perceived stress, with 82% (32/39) experiencing moderate stress and 10% (4/39) experiencing severe stress.

**Figure 2:**
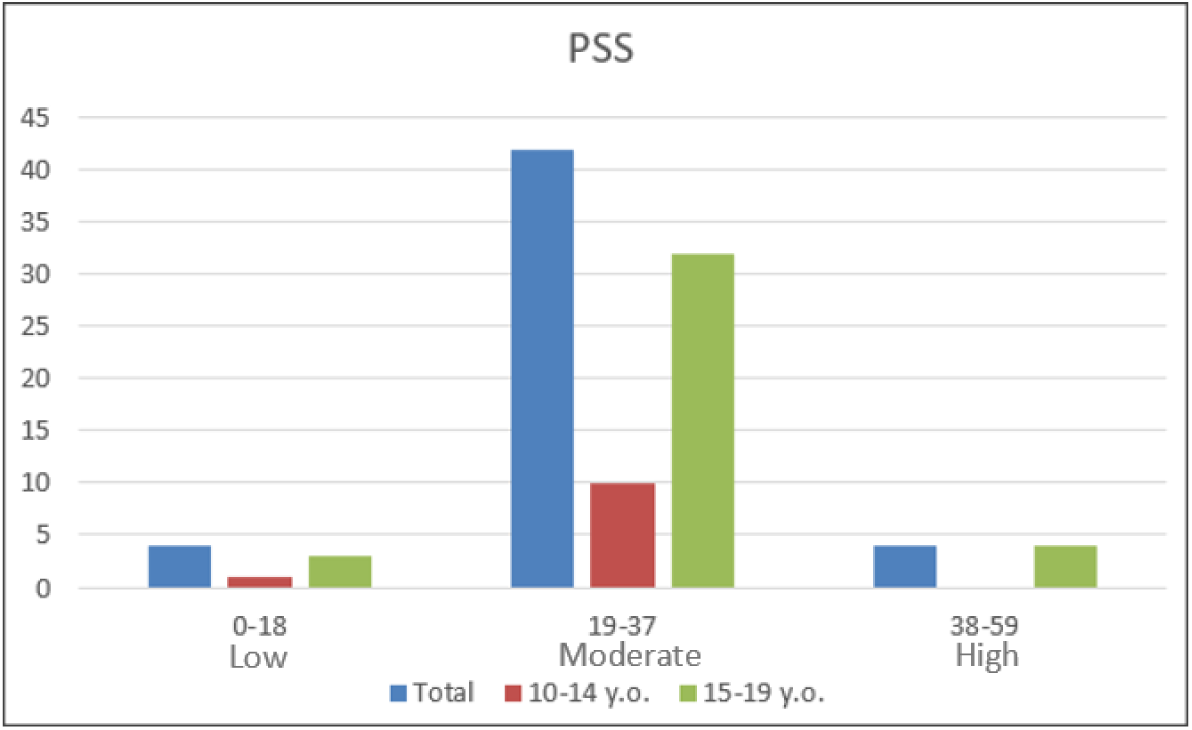
Baseline perceived stress among N=50 ALWH participating in the chatbot study.

Regarding stigma and mental health help-seeking, the overall SSOH mean score was 24 (range: 13-36). By age group, the mean SSOH score was 27 and 23 among 11-to 14-year-olds and 15-to 19-year-olds, respectively.

### Main Outcome

We also used a paired-samples t-test to compare pre-test (mean=6.88, SD=1.85) and post-test (mean=8.12, SD=2.20) scores on the ADQK. After interacting with the chatbot, participants experienced a significant increase in depression knowledge, t(49)= -5.03, p<.001, with a large effect size, d=1.74.

### Chatbot feasibility and acceptability

Table 3 displays feasibility and acceptability ratings for the chatbot. Across the AIM, IAM, and FIM (measuring the chatbot’s acceptability, appropriateness, and feasibility, respectively), total mean scores were ≥3.82/4.00, corresponding to high overall acceptability. Similarly, participants reported high overall intention to use and recommend EVA to friends (overall mean = 10.6/12.0) and satisfaction with EVA’s features and content (range of total mean scores = 4.2-4.66/5.00).

**Table 3:** Endline chatbot feasibility and acceptability among N=50 ALWH participating in the chatbot study.

| Measure | 10-14 years old<br>mean score<br>(range) | 15-19 years old<br>mean score<br>(range) | Total mean score<br>(range) |
| --- | --- | --- | --- |
| **Acceptability of Intervention Measure (AIM)** | 3.91 (0-4) | 3.84 (0-4) | 3.88 (0-4) |
| **Intervention Appropriateness Measure (IAM)** | 3.55 (0-4) | 3.89 (0-4) | 3.82 (0-4) |
| **Feasibility of Intervention Measure (FIM)** | 3.91 (0-4) | 3.87 (0-4) | 3.88 (0-4) |
| **Intention to use and recommend EVA** | 10.09 (3-12) | 10.74 (3-12) | 10.6 (3-12) |
| **Satisfaction with chatbot content and features** | 4.5 (1-5) | 4.46 (1-5) | 4.47 (1-5) |
| **Videos** | 4.27 (3-5) | 4.28 (1-5) | 4.28 (1-5) |
| **Information useful** | 4.54 (3-5) | 4.61 (4-5) | 4.6 (3-5) |
| **Clear and entertaining information** | 4.36 (3-5) | 4.46 (2-5) | 4.44 (2-5) |
| **Entertaining chatbot** | 4.27 (3-5) | 4.17 (1-5) | 4.2 (1-5) |
| **Chatbot easy to use** | 4.27 (4-5) | 4.61 (1-5) | 4.64 (1-5) |
| **Useful self-help skills** | 4.45 (2-5) | 4.48 (2-5) | 4.48 (2-5) |
| **I would ask for professional help through the chatbot.** | 4.27 (3-5) | 4.38 (1-5) | 4.46 (1-5) |
| I really liked the chatbot in general | 4.27 (4-5) | 4.64 (3-5) | 4.66 (3-5) |

### Open-ended text response regarding likes and dislikes of the chatbot

Responses to the open-ended question inviting specific feedback on the chatbot are summarized in Table 4. Participants reported liking the chatbot’s content, visual aids, and ability to connect them with a mental health professional. Dislikes centered on a lack of content personalization, technological limitations, and a lack of human emotional connection.

**Table 4:**
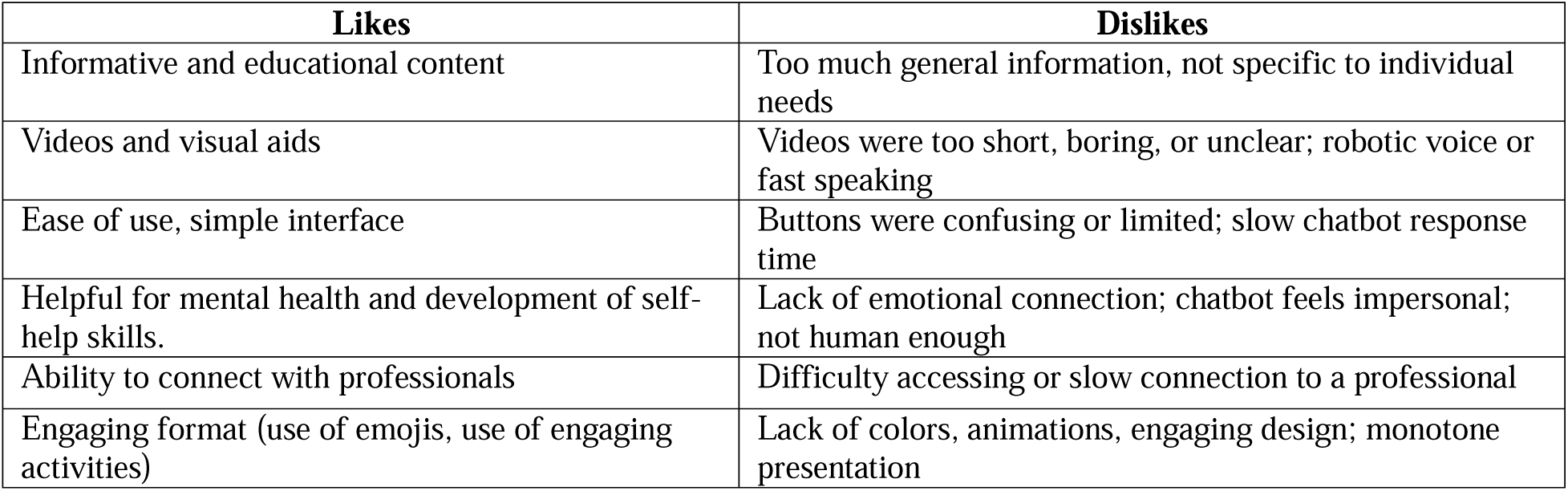
Open-ended text responses on the likes and dislikes of the chatbot.

## Discussion

A single, brief interaction with a novel mental health chatbot resulted in a significant increase in depression knowledge among ALWH in Peru, and the chatbot was rated highly acceptable, appropriate, and feasible by adolescents. Importantly, although study participation was not contingent on mental wellness, 70% and 60% of participants reported current depressive and anxious symptoms, respectively, and 92% of participants had perceived stress levels of moderate or higher. Together, our findings demonstrate the potential positive impact of a mental health chatbot for ALWH and underscore the high levels of mental health symptoms among this population.

The significant increase in depression knowledge following chatbot use (with a large effect size) has important implications. Though psychoeducation alone does not directly treat depressive symptoms, it can increase symptom recognition, reduce stigma, and promote help-seeking behavior [46]. For example, psychoeducation was a core component of a recent multi-session depression and treatment adherence intervention for ALWH in Botswana, which led to improved adherence and lower depressive symptoms [47]. Considering the relatively high reported levels of stigma and mental health help-seeking in the present study, all ALWH should minimally be provided with psychoeducation, which could be easily facilitated by providing access to a chatbot like EVA at the HIV clinic.

Other chatbots exist for people at risk of or living with HIV, including the general population, men who have sex with men, female sex workers, health care workers, and transgender adolescents [48]; however, to our knowledge, EVA is the only chatbot mental health chatbot for ALWH. EVA was rated highly acceptable, feasible, and appropriate by participants in both the 10-14-year-old and 15-19-year-old groups, indicating its appeal across developmental stages. We attribute these findings to the human-centered design approach in partnership with the Youth Advisory Board throughout the chatbot development process, which prioritized ALWH’s content and design preferences from the outset [33]. However, areas for refinement point to design challenges in creating personalized, accessible, and empathic digital health tools. Future versions of EVA could include virtual peer-to-peer support groups and a live telehealth portal for speaking with mental health professionals to increase human interaction.

The EVA chatbot was programmed using software predating the rapid and widespread commercial access to artificial intelligence (AI) and instead followed a predetermined, rules-based format. Many adolescents already access AI chatbots for mental health issues daily: a nationally representative survey conducted among U.S. adolescents and young adults found that a fifth used AI chatbots for mental health advice, and half of those did so monthly [49]. Future versions of EVA could leverage AI to enhance and expand its capabilities to detect participant distress, provide preemptive support to maintain healthy behaviors during such times, and assist youth in understanding how their mental wellness affects ARV adherence. Similarly, AI could be used to tailor the delivery of information and skills based on age, sex, and other relevant variables. Development of AI-enabled tools, however, could be hampered by concerns regarding data safety and confidentiality, a prominent theme we found in previous qualitative research on mental health chatbot acceptability among ALWH and their caregivers [35, 37].

Other concerns that must also be considered include concerns about information bias, fabrications, and other so-called AI “hallucinations” [48] and a mounting evidence base linking the impact of chatbots on adolescent mental health development. A recent review including N=80 studies conducted during 2020-2025, for example, found that while mental health chatbots have demonstrated benefit for adolescents—especially for anxiety, depression, health literacy and health-seeking behaviors—worrisome use, the potential for dependency and risks to social development were recurring themes [50]. Similar findings were reported in a 2025 systematic review and meta-analysis of 26 studies including N=29,637 adolescents and young adults on the effectiveness of AI chatbots on reducing mental distress and promoting health behaviors [51]. In that study, the authors highlight the potential of AI chatbots while also recommending the prioritization of safety protocols and evaluation frameworks. Data on the specific impact of chatbots on Peruvian adolescents’ mental health is lacking; however, more generally, a 2025 study among N=630 high school students found that fear of being without a mobile phone was significantly associated with low self-esteem [52]. Future development of EVA and similar chatbots for adolescents must balance their benefits to mental health with their potential risks.

## Limitations

As a pilot study, our sample size prevented stratification of data by age group, and the results are not generalizable to all ALWH populations. Nonetheless, the sample included ALWH with diverse identities and experiences, including non-heterosexually or homosexually identifying youth; mothers and pregnant girls; and youth who acquired HIV at birth and sexually. Because we recruited participants engaged in HIV care, current antiretroviral use was high, thereby underrepresenting youth with lower antiretroviral adherence who may be experiencing greater mental health morbidity. Nonetheless, overall depression and anxiety symptom prevalence was also high, suggesting that mental health issues are relevant to all youth with HIV, including those currently engaged in HIV care. Finally, this study was an initial step at determining if a chatbot could be used to support ALWH’s mental health and was not designed to measure its impact over time. In addition to a larger, more diverse study sample, future studies should be longitudinal to assess the chatbot’s impact on mental health over time, particularly regarding mental health help-seeking.

## Conclusions

A mental health chatbot developed with and for ALWH increased depression knowledge and was rated highly acceptable and feasible. Further development of mental health chatbots customized for the specific needs of ALWH holds potential as a low-cost, scalable intervention that can both reduce mental health morbidity and support optimal HIV outcomes. Any benefits of increased use of technology for mental health among ALWH must also be weighed against the potential risks they may introduce.

## Data Availability

All data produced in the present study are available upon reasonable request to the authors

## Authors’ contributions

JTG and CC conceived the study, which was further developed and refined by DV, KK and MFF. Chatbot programming was led by DV with support from NP. DV and MT led data collection activities, coordinating participant recruitment with LK. JTG, CC, DV, NP, KK and KG processed and cleaned data. All authors contributed to the final manuscript: JTG drafted the first version, which was reviewed and approved by all authors.

## Acknowledgments

This study emerged as a response to the mental health service gap for adolescents with HIV in Peru and was entirely dependent on the ongoing participation of adolescents with HIV for all aspects of the chatbot development and testing. We are especially grateful for the participation of the Youth Advisory Board in Lima, Peru, for their candid feedback on the chatbot and related study procedures. The authors acknowledge the use of Microsoft Copilot (accessed July 2026) and Grammarly (accessed July 2026) to suggest improvements in this manuscript’s grammar, spelling, clarity and flow. All AI-generated suggestions were reviewed, revised, and approved by the authors, who take full responsibility for the accuracy and integrity of the work.

